# Exploring the ‘Visible Versus Invisible’ Paradigm in Cavernous Sinus Dural Arteriovenous Fistula

**DOI:** 10.1101/2025.02.06.25321830

**Authors:** Jingwei Zheng, Shixing Su, Hua Lu, Sheng Liu, Shengjun Zhou, Qi Jia, Xiang Bao, Zhenqiang Li, Haihang Zhou, Guoqiang Zhang, Zhijie Jiang, Fei Liu, Si Hu, Zixin Wang, Jun Yu, Liang Xu

**Affiliations:** Department of Neurosurgery, Second Affiliated Hospital, School of Medicine, Zhejiang University, Hangzhou, Zhejiang, China; Clinical Research Center for Neurological Diseases of Zhejiang Province, Hangzhou, China; Neurosurgery Center, Department of Cerebrovascular Surgery, Engineering Technology Research Center of Education Ministry of China on Diagnosis and Treatment of Cerebrovascular Disease, Zhujiang Hospital of Southern Medical University, Guangzhou, Guangdong, China; Department of Neurosurgery, Jiangsu Province Hospital and Nanjing Medical University First Affiliated Hospital, Nanjing, Jiangsu, China; Department of Interventional Radiology, The First Affiliated Hospital of Nanjing Medical University, Nanjing, China; Department of Neurosurgery, The First Affiliated Hospital of Ningbo University, Liuting Street 59, Ningbo, 315010, Zhejiang, China; Department of Neurosurgery, Affiliated Hospital of Nantong University, Nantong, China; Department of Neurosurgery, Jinhua Municipal Central Hospital, Jinhua, Zhejiang, China; Department of Neurosurgery, Ningbo Medical Center Lihuili Hospital, Ningbo, Zhejiang, China; Department of Neurosurgery, The Second Affiliated Hospital of Jiaxing University, Jiaxing, China; Department of Neurosurgery, Affiliated Huzhou FuYin Hospital of Huzhou University, Huzhou, Zhejiang, China

## Abstract

**Background:** Cavernous sinus dural arteriovenous fistulas (CS-DAVFs) present significant treatment challenges when the inferior petrosal sinus (IPS) is not opacified during cerebral angiography. However, the widely accepted transvenous IPS recanalization approach is associated with a high failure rate. The consistently visible superior ophthalmic vein (SOV) offers a promising alternative, though it has yet to be fully evaluated in large-scale studies.

**Methods:** This retrospective, case-control study was conducted between May 2017 and October 2024. Data collection for this multicenter, population-based study took place across eight tertiary referral centers. Eligible patients were diagnosed with CS- DAVF with occluded IPS. Endovascular treatment via the transvenous SOV approach versus the IPS recanalization approach in patients with occluded IPS.

**Results:** Of 178 eligible cases, 70 cases (39.3%) were treated using the transvenous SOV approach, while 108 cases (60.7%) underwent the transvenous IPS approach. The initial treatment success rate was significantly higher in the SOV group compared to the IPS group (91.4% vs. 75.9%; odds ratio [OR], 3.38; 95% CI, 1.30–8.35; *P* = 0.0092). The overall complication rate was 1.4% in the SOV group and 2.8% in the IPS group (OR, 0.51; 95% CI, 0.04–3.47; *P* > 0.9999). After classifying the SOV approach into simple and complex types, the SOV-simple type further demonstrated significant advantages, including shorter average operation times (126.20 ± 46.99 minutes, *P* = 0.0197) and a higher initial treatment success rate (95.7%, *P* = 0.0027) compared to the IPS group.

**Conclusion:** The SOV approach should be considered a first-line treatment for CS- DAVF patients with ‘invisible’ IPS. These findings establish a new treatment standard, underscoring the importance of precise preoperative classification and individualized surgical planning.

## Introduction

A cavernous sinus dural arteriovenous fistula (CS-DAVF) is an abnormal arteriovenous shunt involving the dura mater surrounding the cavernous sinus (CS) ^1,2^. The clinical manifestations of CS-DAVF are largely determined by its venous drainage patterns. The clinical manifestations of CS-DAVFs are largely determined by their venous drainage patterns. Common initial symptoms include ophthalmological issues such as proptosis and diplopia, primarily due to fistula drainage via the superior ophthalmic vein (SOV) in most cases ^2^. Timely intervention is often necessary to alleviate these intractable ocular complications.

Endovascular embolization via the inferior petrosal sinus (IPS) is widely regarded as the first-line treatment for CS-DAVF ^3^. The IPS offers a direct and accessible route to the CS, making it the preferred approach for embolization ^4^. However, the endovascular treatment via IPS approach can sometimes pose challenges, particularly in cases of IPS occlusion, hypoplasia, or absent venous communication between the IPS and the cavernous sinus fistula ^3^. The current reliance on IPS recanalization is primarily based on limited data from single-center studies and expert consensus. Both prior studies and our own single-center experience highlight a notable rate of unsuccessful IPS access ^3,4^, alongside a significant risk of complications, including subarachnoid hemorrhage and cranial nerve palsy ^5–7^.

Therefore, there remains significant potential for further enhancement of existing standard strategies. Given the predictable nature of venous drainage, the SOV is consistently visible, offering a viable alternative in these cases ^6^. However, its safety and efficacy remain insufficiently evaluated in large-scale study. To address this gap, the multicenter retrospective case-control study was conducted to compare the efficacy and safety of the visible SOV approach with the ‘invisible’ IPS approach in CS-DAVF patients with an occluded IPS. This study aims to provide robust evidence to guide clinical decision-making and improve outcomes in this complex patient population.

## Methods

### Study Design and Setting

This multicenter, retrospective, case-control study was conducted across eight high-volume tertiary care centers specializing in neurovascular interventions between May 2017 and October 2024. The study aimed to compare the safety and efficacy of the transfemoral vein-SOV approach with the IPS recanalization approach in treating patients with CS-DAVF and IPS occlusion. The study adhered to the principles of the Declaration of Helsinki and received ethical approval from the institutional review boards of all participating centers. Given its retrospective nature, the requirement for informed consent was waived. The reporting followed the Strengthening the Reporting of Observational Studies in Epidemiology (STROBE) guideline.

### Participants

A total of 245 consecutive cases with angiographically confirmed CS-DAVFs who underwent transvenous endovascular treatment during the study period were screened. After applying inclusion and exclusion criteria, 178 cases were eligible for analysis. Among these, 70 cases treated using the FV-SOV or superficial temporal vein (STV)-SOV approach constituted the case group, while 108 cases treated using the IPS approach served as the control group. The specific inclusion and exclusion criteria, angiographic evaluation, endovascular procedure, outcomes measures, follow-up and statistical analysis are detailed in the **Supplement Material.**

## Results

### Procedural Results

Out of 245 CS-DAVF cases treated with transvenous embolization at all participating centers during the study period, 67 cases with a ‘visible’ IPS were excluded (**Figure 1A**). As a result, 178 cases with an ‘invisible’ IPS were included in the analysis. Among these, 70 patients (39.3%) were treated using the transvenous SOV approach, while 108 patients (60.7%) underwent the transvenous IPS approach (**Figure 1B**).

**Figure 1.**
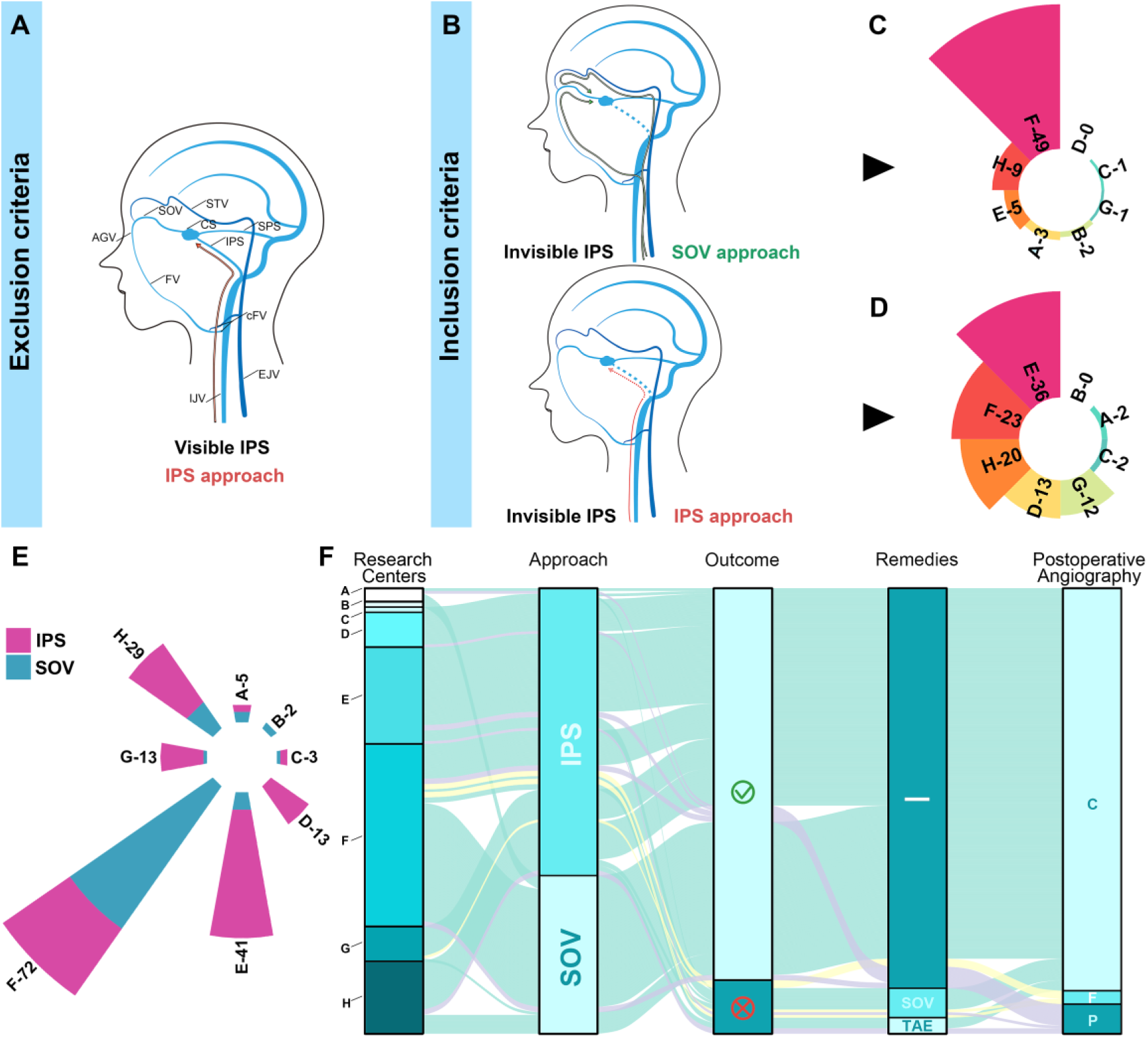
The Enrollment and Treatment Procedure Of CS-DAVF Patients. **A.** Patients with patent IPS during angiography were excluded. **B.** Patients with occluded IPS during angiography were included. **C.** The rose diagram illustrates the distribution of patients treated with transvenous SOV approach across all eight participating centers. **D.** The rose diagram illustrates the distribution of patients treated with transvenous IPS approach across all eight participating centers. **E.** The rose diagram illustrates the distribution of patients treated with both transvenous SOV and IPS approach across all eight participating centers. **F.** The Sankey diagram depicts the progression of treatment (IPS or SOV), initial treatment outcomes (success or failure), remedies (NA, SOV or TAE) and the final immediate postoperative angiography results (C: complete embolization; F: failure of operation; P: partial embolization) for each case. AGV: angular vein; SOV: superior ophthalmic vein; FV: facial vein; cFV: common facial vein; CS: cavernous sinus; STV: superficial temporal artery; IPS: inferior petrosal sinus; SPS: superior petrosal sinus; EJV: external jugular vein; IJV: internal jugular vein.

The circular histogram (rose diagram, **Figures 1C–E**) illustrates the distribution of cases across the eight participating centers. Center ‘F’ contributed the largest number of cases treated with the transvenous IPS-SOV approach, while Center ‘E’ had the highest number of cases treated with the transvenous IPS recanalization approach. Notably, Center ‘F’ also enrolled the highest total number of cases.

The Sankey diagram (**Figure 1F**) further visualizes the progression of treatment and outcomes for each case. Of the total cases, 108 patients were initially treated with the transvenous IPS approach. However, in 20 of these cases, the microcatheter and microguidewire were unable to advance into the CS near the fistula. Consequently, three patients opted out of further treatment, six patients underwent trans-arterial endovascular treatment (TAE) as a remedial measure, and 11 patients were switched to the transvenous SOV approach. The remaining 59 patients, initially treated with the transvenous SOV approach, did not require any further intervention. Ultimately, endovascular treatment failed in five patients (Excluding patients with initial interventional failure), partial embolization was achieved in 11 patients, and complete embolization was accomplished in 151 patients.

### Baseline Characteristics and Endovascular Treatment Outcomes

The detailed characteristics are summarized in **Table S1**. The median age was 58 years (range, 13–81) in the SOV group and 61 years (range, 15–83) in the IPS group. The cohort consisted of 122 females (68.5%) and 56 males (31.5%) (**Figure 2A**). Consistent with the previous findings (5), 81.8% of patients (54/66, excluding four with invisible FVs) exhibited fistula vessels (FVs) draining into the external jugular vein (EJV) rather than the internal jugular vein (IJV), representing a significant deviation from the typical vascular anatomy (**Figure 2A**).

**Figure 2.**
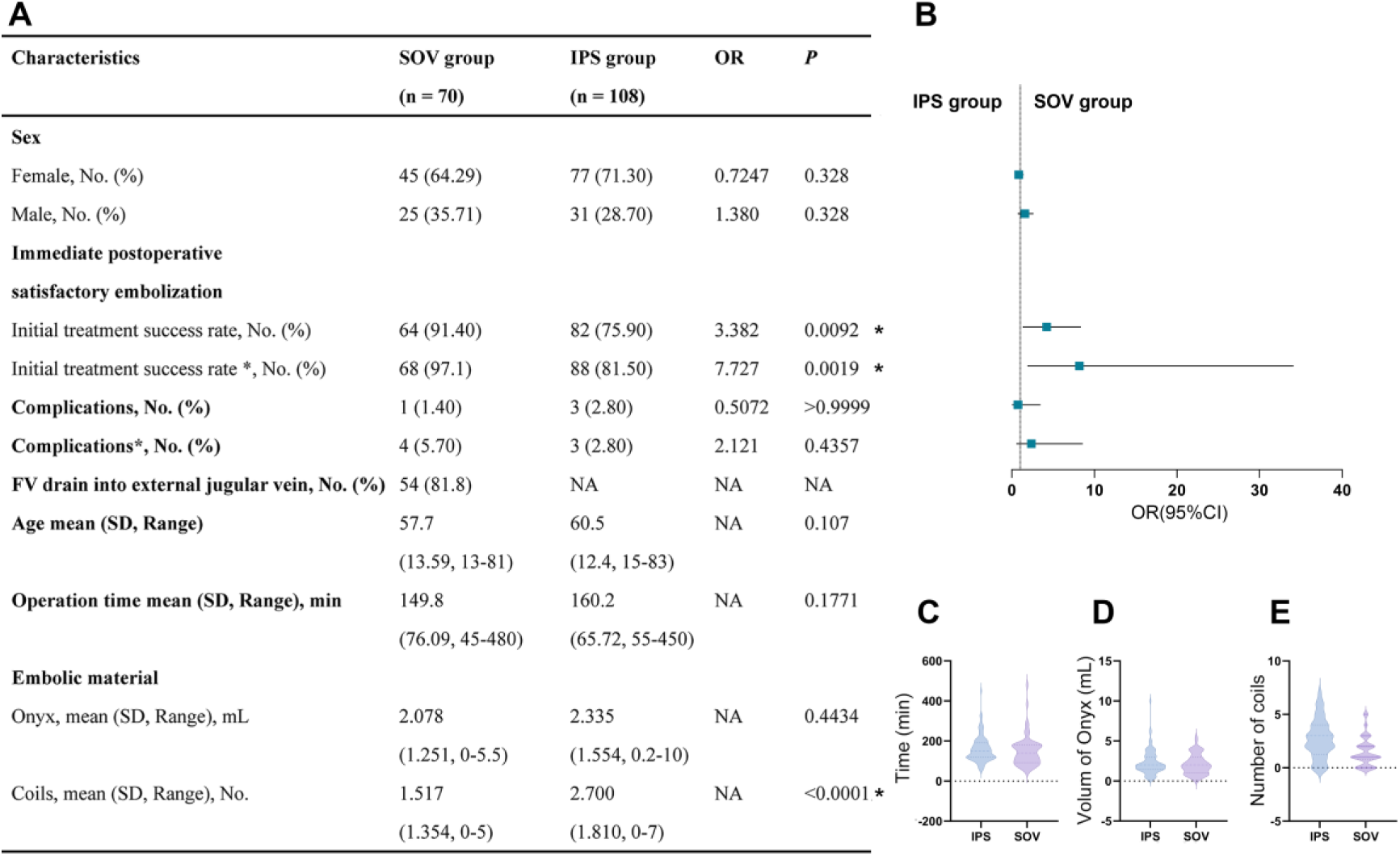
The Baseline Characteristics and Endovascular Treatment Outcomes. **A.** Baseline characteristics of recruited patients. Initial treatment success rate: the rate of immediate postoperative complete embolization Initial treatment success rate* (adjusted initial treatment success rate): the rate of immediate postoperative complete embolization and near-complete embolization. Complication: permanent or long-term complication including cranial nerve palsy, hemorrhage and so on. Complication*(adjusted complication rate): transient or long-term or permanent complication including cranial nerve palsy, hemorrhage and so on. **B.** The forest plots. **C.** The violin plot of operation time. **D.** The violin plot of volume of Onyx. **E.** The violin plot of number of coils. SOV: superior ophthalmic vein; IPS: inferior petrosal sinus.

#### Primary Outcomes

The initial treatment success rate was 91.4% (64/70) in the SOV group and 75.9% (82/108) in the IPS group (OR, 3.38; 95% CI, 1.30–8.35; *P* = 0.0092; **Figure 2B**). Similarly, the adjusted success rate was 97.1% (68/70) in the SOV group and 81.5% (88/108) in the IPS group (OR, 7.73; 95% CI, 1.88–34.15; *P* = 0.0019; **Figure 2B**).

In the SOV group, one patient developed long-term mild tinnitus, and three patients experienced transient postoperative complications, including 3rd cranial nerve palsy (n=2) and 6th cranial nerve palsy (n=1). In the IPS group, two patients developed long-term 3rd cranial nerve palsy, and in one case, the microcatheter could not be removed and was ultimately left in the vessel. The overall complication rate was 1.4% (1/70) in the SOV group and 2.8% (3/108) in the IPS group (OR, 0.51; 95% CI, 0.04–3.47; *P* > 0.9999; **Figure 2B**). The adjusted complication rate was 5.7% (4/70) in the SOV group and 2.8% (3/108) in the IPS group (OR, 2.12; 95% CI, 0.55– 8.59; *P* = 0.4357; **Figure 2B**).

#### Secondary Outcomes

The average operation time was 149.80 ± 76.09 minutes in the SOV group and 160.2 ± 65.72 minutes in the IPS group (*P* = 0.1771; **Figure 2A, C**). There was no significant difference in the amount of Onyx used between these groups (*P* = 0.4143; **Figure 2A, D**), with the SOV group using 2.078 ± 1.251 mL and the IPS group using 2.335 ± 1.554 mL. However, the number of coils used differed significantly between the groups: the SOV group used 1.517 ± 1.354 coils, while the IPS group used 2.70 ± 1.81 coils (*P* < 0.0001; **Figure 2A, E**). Notably, the total medical costs for embolic materials (coils and Onyx) were significantly higher in the IPS group compared to the SOV group (**Figure S1**, *P* = 0.0001).

### Angiographic Classifications for Treatment Options

#### Simple Type

Type I (**Figure 3A**): The junction of the SOV and FV was relatively smooth (red circle), without hairpin curves, allowing the microcatheter to be successfully advanced into the CS via the SOV route. The FV opening also showed no large angulation (red circle), enabling easy catheterization of the support catheter (4Fr angiographic or 5Fr guiding catheter) into the FV, providing sufficient support for the microcatheter.

**Figure 3.**
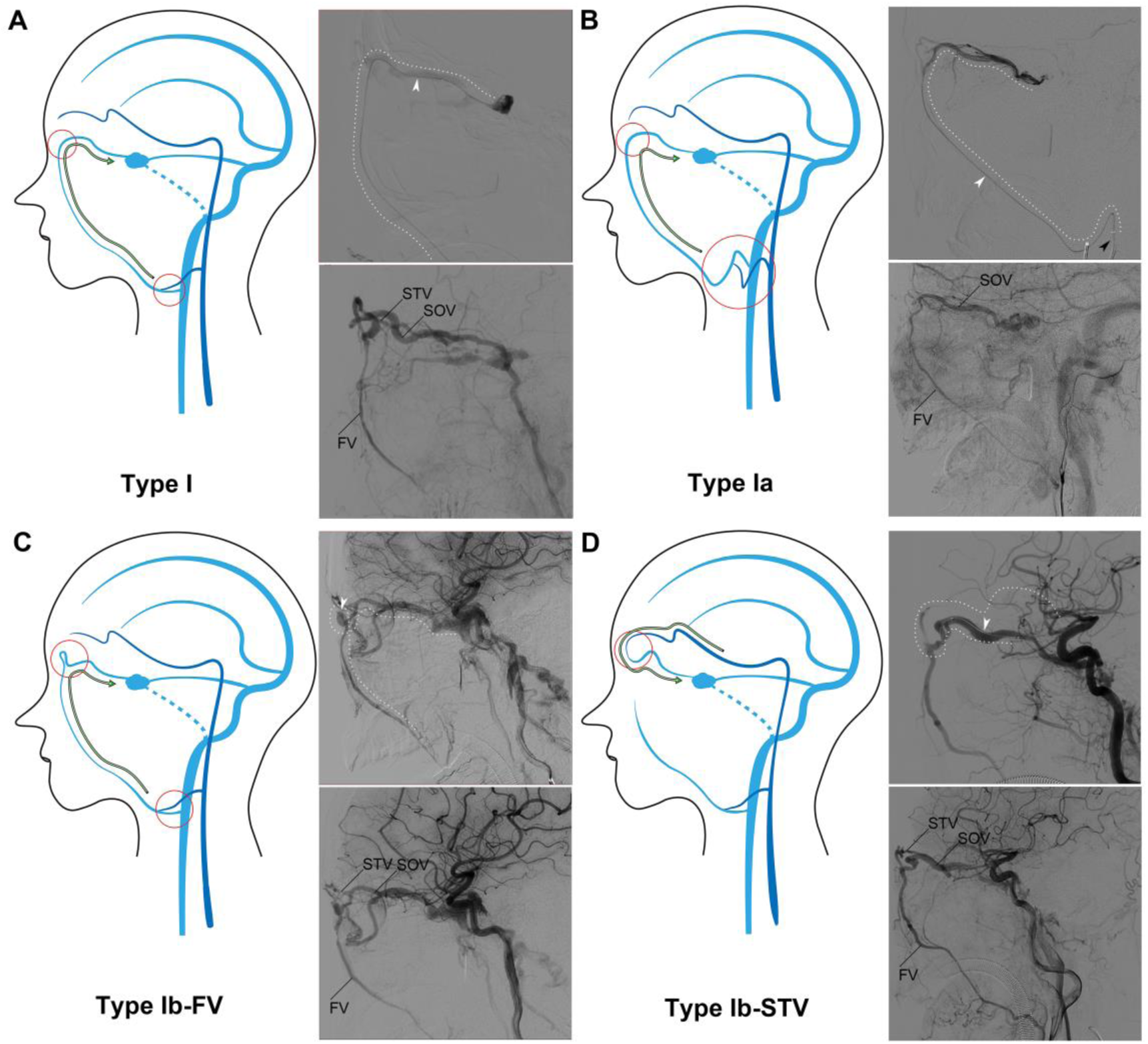
The classification of Simple Type SOV. **A.** Type I; **B.** Type Ia; **C.** Type Ib-FV; **D.** Type Ib-STV. The dotted line indicates the path of the microcatheter (white arrow), the black arrow indicates the support catheter. SOV: superior ophthalmic vein; FV: facial vein; STV: superficial temporal artery; IPS: inferior petrosal sinus.

Type Ia (**Figure 3B**): The junction of the SOV and FV was smooth, with no hairpin curve. However, the FV opening had a large angulation (red circle), making support catheter insertion difficult (black arrow). Despite the lack of proximal support, the smooth distal junction between the SOV and FV allowed successful advancement of microcatheter.

Type Ib-FV (**Figure 3C**): Though the SOV-FV junction had a hairpin curve, the smooth opening of FV made the catheterization of the support catheter easy. Adequate proximal support ensured successful microcatheter advancement through the distal hairpin curve.

Type Ib-STV (**Figure 3D**): Generally, the STV was not recommended as the primary access route due to the reverse angle at the junction of the STV and SOV. However, in the absence of a hairpin curve at this junction, this approach may still be feasible for microcatheter catheterization.

#### Complex Type

Type II (**Figure 4A)**: The smooth opening of the FV facilitated easy catheterization of the support catheter, which provided sufficient proximal support for the microcatheter. However, the presence of multiple hairpin curves (≥ 2) at the SOV- FV junction complicated the delivery of the microcatheter into the CS near the fistula shunt point.

**Figure 4.**
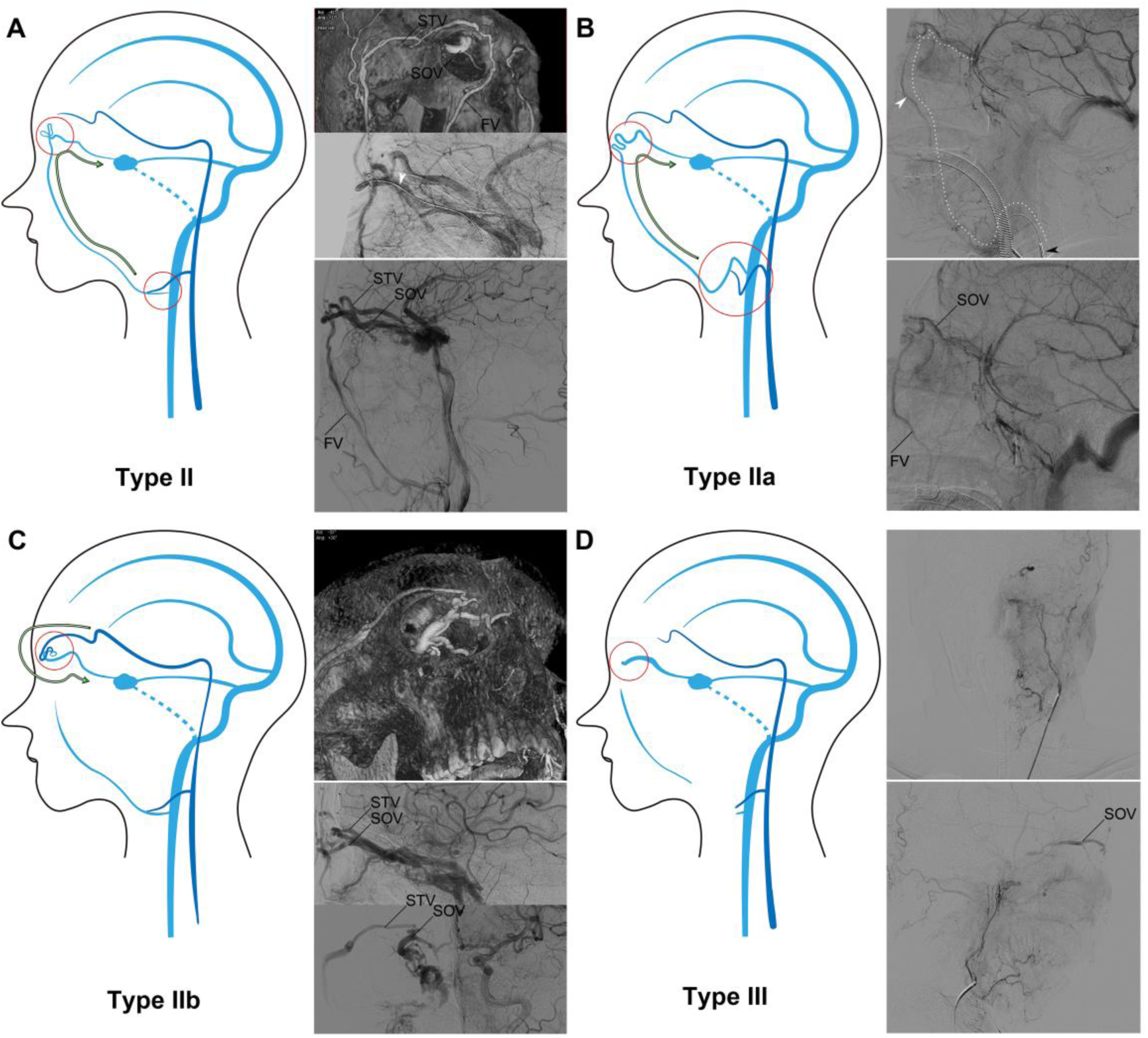
The classification of Complex Type SOV. **A.** Type II; **B.** Type IIa; **C.** Type IIb; **D.** Type III. The dotted line indicates the path of the microcatheter (white arrow), the black arrow indicates the support catheter. SOV: superior ophthalmic vein; FV: facial vein; STV: superficial temporal artery; IPS: inferior petrosal sinus.

Type IIa (**Figure 4B**): The FV opening exhibited a large angulation, making support catheter insertion challenging (black arrow). In the absence of adequate proximal support, even a single distal hairpin curve (≥ 1) at the SOV-FV junction can hinder microcatheter navigation.

Type IIb (**Figure 4C**): Typically, the STV formed a reverse angle at the STV- SOV junction. The presence of even a single distal hairpin curve (≥ 1) at the SOV-FV junction could significantly hinder the advancement of the microcatheter.

Type III (**Figure 4D**): These approaches to the SOV, including the FV, STV, and IPS, were not visible during angiography, which made navigation and catheterization more challenging.

### Clinical Outcomes Based on Angiographic Classifications

According to the anatomical characteristics observed on angiography and the difficulty associated with interventional treatment (**Figure 5A, B**), the SOV approach was divided into two categories: simple type (Type I, Type Ia, Type Ib-FV, Type Ib- STV) and complex type (Type II, Type IIa, Type IIb, Type III).

**Figure 5.**
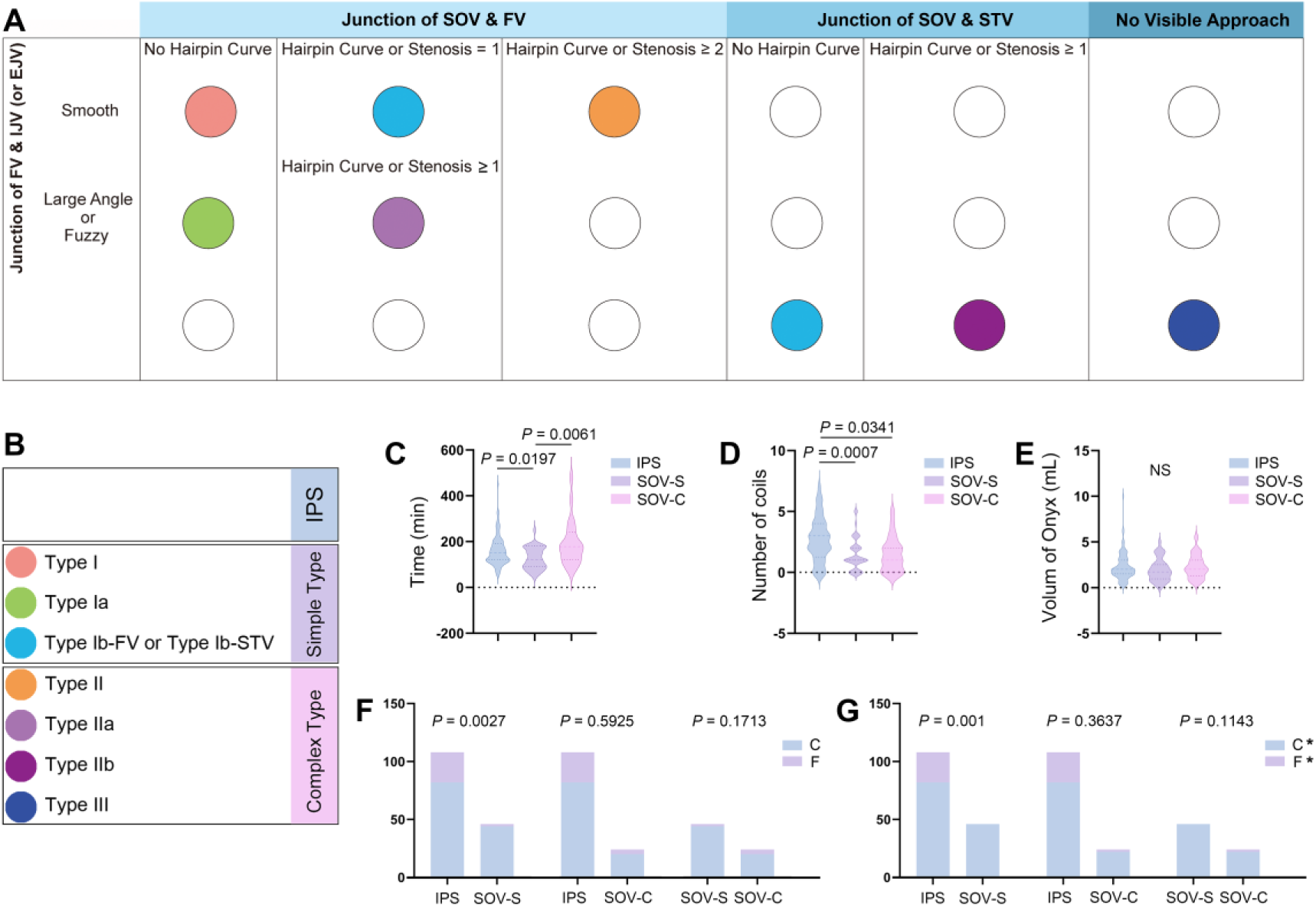
Endovascular treatment Outcomes after SOV classification. **A & B.** SOV classification reference table **C.** The violin plot of operation time. **D.** The violin plot of number of coils. **E.** The violin plot of volume of Onyx. **F.** The histogram of initial treatment success rate (the rate of immediate postoperative complete embolization). **G.** The histogram of adjusted initial treatment success rate, the rate of immediate postoperative complete embolization and near-complete embolization. SOV: superior ophthalmic vein; FV: facial vein; STV: superficial temporal artery; IPS: inferior petrosal sinus; SOV-S: SOV simple type; SOV-C: SOV complex type.

Inconsistent with the results before the classification, our analysis revealed that the average operation time was significantly shorter in the SOV-S group (126.20 ± 46.99 minutes) compared to the IPS group (160.2 ± 65.72 minutes; *P* = 0.0197; **Figure 5C**). Furthermore, procedures in the SOV-C group took more time (191.8 ± 98.31 minutes) than in the SOV-S group (*P* = 0.0061; **Figure 5C**). However, there was no significant difference in the average operation time between the IPS group and the SOV-C group (*P* = 0.6129; **Figure 5C**).

The number of coils used varied significantly between groups, with the SOV-S group using 1.487 ± 1.335 coils (*P* = 0.0007; **Figure 5D**) and the SOV-C group using 1.600 ± 1.392 coils (*P* = 0.0341; **Figure 5D**), compared to the IPS group (2.700 ± 1.810 coils; **Figure 5D**). However, there was no significant difference in the amount of Onyx used across groups (**Figure 5E**), with the IPS group using 2.335 ± 1.554 mL, the SOV-S group using 1.947 ± 1.225 mL, and the SOV-C group using 2.325 ± 1.292 mL. Similarly, as demonstrated in **Figure S2**, the total medical costs for embolic materials (coils and Onyx) were significantly higher in the IPS group compared to the SOV-S group (*P* = 0.0025) and the SOV-C group (*P* = 0.0336).

The SOV-S group demonstrated a significantly higher initial treatment success rate compared to the IPS group (95.7%, 44/46 vs. 75.9%, 82/108; OR, 6.976; 95% CI, 1.728–30.73; P = 0.0027; **Figure 5F**). Similarly, the adjusted success rate was 100% (46/46) in the SOV-S group compared to 81.5% (88/108) in the IPS group (OR, ∞; 95% CI, 2.764–∞; *P* = 0.001; **Figure 5F**). Intergroup comparisons showed no significant difference among other groups.

## Discussion

The primary objective of CS-DAVF management is to eliminate both subarachnoid and SOV drainage. Nowadays, the endovascular treatment for CS- DAVF has evolved from initial trans-arterial embolization to the widely accepted transvenous IPS approach embolization. However, in a significant proportion of patients, the IPS is not opacified on cerebral angiography. Although historically challenging, navigating the microcatheter to the fistula in CS-DAVF cases with “invisible” IPS has predominantly relied on IPS recanalization as the preferred treatment option. Notably, previous studies highlight a notable rate of unsuccessful IPS recanalization ranging from 54.3% ^4^ to 93% ^6^. Therefore, there is considerable potential for further optimization of current standard strategies.

Recently, the trans-SOV approach has emerged as an alternative therapy for failed IPS recanalization. Unlike the angiographically “invisible” IPS, the “visible” SOV drainage provides a reliable roadmap for microcatheter navigation. A few studies have demonstrated that the transvenous SOV approach may offer superior efficacy and safety compared to IPS recanalization ^3,4,6^. Previous single-center study also showed promising results with a 97% immediate complete embolization rate, a 100% long-term symptom resolution rate ^7^. Therefore, it arouses sparking debate that whether the transfemoral SOV route may be a viable alternative approach, or even should be prioritized over IPS recanalization. In the present study, SOV approach offers distinct advantages over the traditional IPS recanalization approach, demonstrating significantly higher initial treatment success rates (91.4% vs. 75.9%). Notably, successful IPS recanalization often depends on fortuitous anatomical conditions. Particularly in cases of IPS hypoplasia, attempts to recanalize the IPS are inevitably destined to fail. By contrast, the clearly identifiable SOV provides a more reliable and precise pathway compared to the IPS, which may not be visible on cerebral angiography.

The SOV approach has historically been met with skepticism, largely due to its perceived complexity and extended procedural distance. The difficulty of the SOV route is often considered to correlate with longer procedural time. However, our findings revealed that the SOV approach did not significantly prolong procedural duration compared with IPS recanalization, challenging previously held assumptions. Despite the SOV being more easily visualized, catheterization can be challenging or even unsuccessful due to tortuosity or stenosis of the supraorbital, angular, and common FVs. Moreover, improper manipulation may lead to rupture of the SOV and intraorbital hemorrhage, potentially resulting in vision loss. Considering the inherent surgical risks and the challenges, the procedure can be particularly daunting for neurosurgeons. This underscores the rationale for proposing a systematic classification.

In the present study, we are the first to classify SOV anastomoses into two types: simple (**Figure 3**) and complex (**Figure 4**), highlighting how anatomical variations impact the feasibility of endovascular treatment. The proposed classification primarily focuses on two critical challenges in establishing an interventional access route: the ability of microdevices to pass through the FV opening and the junction of FV-SOV or STV-SOV. These factors play a pivotal role in determining the success of endovascular procedures and surgical outcomes. Such classification offers a structured framework for preoperative evaluation, enabling surgeons to accurately assess risks and difficulties, thereby facilitating tailored surgical strategies.

Our findings underscore the procedural efficiency of the simple type SOV approach, characterized by shorter operation times, reduced use of embolic materials (coils), and lower medical costs compared with IPS recanalization. Even in cases with SOV-complex type, operation times were comparable to the IPS group, while retaining the advantages of minimized embolic material usage and reduced overall medical expenses.

For neurosurgeons inexperienced with the SOV approach, we recommend initially focusing on simple SOV anatomy, particularly the FV-SOV or STV-SOV junctions. Accurate identification of simple SOV characteristics is crucial for optimizing the interventional approach. It is worth noting that both simple and complex SOV features can coexist in the same patient. For example, in cases illustrated in **Figure S3**, a complex STV-SOV route (Type IIb) initially failed due to tortuosity; however, switching to the simple FV-SOV route (Type I) allowed successful advancement of microdevices into the CS. This highlights the importance of precise SOV classification in selecting the optimal surgical approach.

Notably, even the SOV-complex type demonstrates distinct advantages over the IPS recanalization route, albeit with a necessary learning curve to achieve proficiency. To address the challenges associated with SOV-complex anatomy, we propose the following recommendations:

1. Wire-Loop Technique: This method is invaluable for advancing microdevices through tortuous venous anatomy while minimizing the risk of venous rupture.
2. External Pressure Application: Targeted external pressure can improve system stability or redirect microguidewires to facilitate passage through difficult anatomy.

Detailed demonstrations of these techniques are provided in the **Supplementary Video**. Mastery of these methods enables neurosurgeons to navigate the micro- guidewire with enhanced precision and safety, ensuring successful progression toward the shunt point.

Regarding the amount of embolic materials used and associated medical costs, our experience suggests that early opacification of the SOV during cerebral angiography reliably indicates the fistula shunt point at the confluence of the SOV and CS ^7^. In such scenarios, full occlusion of the CS is unnecessary. By leveraging this anatomical characteristic, we have consistently achieved precise embolization while mitigating the risks associated with overpacking. Our preference for combining Onyx and detachable coils leverages their complementary properties, forming a ‘reinforced concrete structure’. This synergy allows for precise control of Onyx within the designated target zone (the shunt point), ensuring effective embolization while reducing the likelihood of Onyx reflux. We believe this approach strikes the best balance between efficacy, safety and cost-effectiveness.

An intriguing finding in the present study is that 81.8% of patients exhibited FVs draining into the EJV rather than the IJV, revealing a notable anatomical deviation from classic vascular anatomy. This raises the question of whether this variation is specific to East Asian populations or represents a characteristic feature of FVs in CS- dAVF patients. Interestingly, in patients undergoing carotid endarterectomy, the majority of FVs continued to drain into the IJV, suggesting that CS-dAVF may induce alterations in FV drainage patterns. However, it remains unclear whether these altered drainage pathways contribute to the development of CS-dAVF or are a consequence of the condition itself.

### Limitations

Selection bias may have occurred due to the non-randomized nature of the cohort. Additionally, variations in treatment protocols could impact the generalizability of the findings.

## Conclusions

In conclusion, the SOV approach should not be regarded merely as an alternative but rather as a first-line treatment for CS-DAVF patients with “invisible” IPS. Especially for patients classified with simple SOV anatomy, the SOV approach should be prioritized over IPS recanalization. These findings establish a new standard of care for this patient population, emphasizing the need for precise preoperative classification and tailored surgical planning.

## Data Availability

All raw data used in this manuscript are available on reasonable request.

## Acknowledgments

None

---

**Figure S1. The total medical costs for embolic materials**

**IPS group versus SOV group, SOV: superior ophthalmic vein; IPS: inferior petrosal sinus.**

**Figure S2. The total medical costs for embolic materials**

**IPS group versus SOV-S group versus SOV-C group, SOV: superior ophthalmic vein; IPS: inferior petrosal sinus; SOV-S: SOV simple type; SOV-C: SOV complex type.**

**Figure S3. An representive case with both simple and complex SOV type**

**The complex STV-SOV route (Type IIb) initially failed due to tortuosity; The simple FV-SOV route (Type I) allowed successful advancement of microdevices into the CS. The dotted line indicates the path of the microcatheter (white arrow), the white circle indicates the shunt point at the CS.**

**Supplementary Video. Application cases of wire-loop technique**

## References

1. Gandhi D, Chen J, Pearl M, Huang J, Gemmete JJ, Kathuria S. Intracranial dural arteriovenous fistulas: classification, imaging findings, and treatment. AJNR Am J Neuroradiol. 2012;33:1007–1013. doi: 10.3174/ajnr.A2798

2. Ali MH, Jones S, Moss HE. Unilateral Proptosis, Redness, Diplopia, and Numbness in a Young Woman. JAMA Ophthalmol. 2016;134:1325–1326. doi: 10.1001/jamaophthalmol.2016.2129

3. Wenderoth J. Novel approaches to access and treatment of cavernous sinus dural arteriovenous fistula (CS-DAVF): case series and review of the literature. J Neurointerv Surg. 2017;9:290–296. doi: 10.1136/neurintsurg-2016-012742

4. Rhim JK, Cho YD, Park JJ, Jeon JP, Kang HS, Kim JE, Cho WS, Han MH. Endovascular Treatment of Cavernous Sinus Dural Arteriovenous Fistula With Ipsilateral Inferior Petrosal Sinus Occlusion: A Single-Center Experience. Neurosurgery. 2015;77:192–199; discussion 199. doi: 10.1227/NEU.0000000000000751

5. Kirsch M, Henkes H, Liebig T, Weber W, Esser J, Golik S, Kuhne D. Endovascular management of dural carotid-cavernous sinus fistulas in 141 patients. Neuroradiology. 2006;48:486–490. doi: 10.1007/s00234-006-0089-9

6. Fujita A, Kohta M, Sasayama T, Kohmura E. Impact of transvenous embolization via superior ophthalmic vein on reducing the total number of coils used for patients with cavernous sinus dural arteriovenous fistula. Neurosurg Rev. 2021;44:401–409. doi: 10.1007/s10143-019-01227-9

7. Xu L, Zheng J, Ling C, Chen X, Fang B, Qian C, Xu J, Yu J. ’An eye for an eye’ therapeutic strategy for cavernous sinus dural arteriovenous fistula: a single-center experience. J Neurointerv Surg. 2024. doi: 10.1136/jnis-2023-021343

